# Attitude towards Mental Help-Seeking, Motivation, and Economic Resources in Connection with Positive, Negative, and General Psychopathological Symptoms of Schizophrenia: A Pilot Study of a Psychoeducation Program

**DOI:** 10.1101/2022.07.01.22277148

**Authors:** Qasir Abbas, Khawar Bilal Baig, Urooj Sadiq, Muhammad Umar Khan, Mafia Shahzadi, Zoobia Ramzan

## Abstract

**Background:** Pharmacological treatment is usually the first line of treatment for schizophrenia, but more strategies are needed to augment this treatment to promote better outcomes. It is known that adherence to pharmacological treatment in schizophrenia patients can be increased by working with their insight into their disorder. In literature, many programs have been found to increase mental help-seeking and reducing symptom severity but most are from the Western cultures and/or are conducted with people attending any institution (i.e., a university or an in-patient care unit) and with specific age ranges (i.e., young adults or older adults). However, in the noninstitutionalized population of different age groups of Pakistan, there is a need to find ways (alongside medication) that promote attitude toward mental help-seeking and reduce symptom severity. Therefore, the current pilot study was designed to investigate the impact of a psychoeducation program on increasing patients’ motivations and help-seeking attitudes toward treatment, reducing the severity of the symptoms, and the role of financial sources in the course of their illness.

**Methodology:** In this pilot study, we targeted diagnosed patients with schizophrenia disorder from different hospitals and primary care clinics. After eligibility screening, 255 participants were included, and 220 completed the psychoeducation program. Both men, 143(56.08%) and women, 112(43.82%) with marital statuses of being single 123(48.24%), married 98(38.43%) and divorced/widower/widowed 34(13.33%) were included. Respondents’ age range was 18-52 years (M=35.45, SD=10.27).

**Results:** Findings revealed that significant change in symptoms severity was observed after 16-weeks psycho-education program on positive symptoms (Md=21.05, n=220) compared to before (Md=25.00, n=220, z=-12.47, p=.000, η_p_^2^= .59, negative symptoms (Md=15.74, n=220) compared to before (Md=17.44, n=220, z=-9.52, p=.000, η_p_ ^2^= .45, and general psychopathological symptoms (Md=38.32, n=220) compared to before (Md=43.40, n=220, z=-12.72, p=.000, η_p_^2^= .61. Similarly, on HSAT (Md=39.03, n=220) compared to before (Md=28.27, n=220, z=-10.43, p=.000, η_p_ ^2^= .50, and PMFT (Md=5.69, n=220) compared to before (Md=4.85, n=220, z=-12.43, p=.000, η_p_ ^2^= .59 respectively. Change in patients’ motivation after 16-weeks at low motivation level was -55(25%) (this category got reduced as people moved to better motivation levels), at moderate motivation level it was 10(4.55%) and at high motivation level it was 45(20.45%). Symptoms severity reduced in across all income groups but patients in low-income group tended to gain more from the psychoeducation programas compared to middle- and high-income group in both pre and post treatment.

**Conclusion:** It is concluded that our psychoeducation program helps promote patients’ motivation and help-seeking attitude toward treatment, and helps reduces positive, negative, and general symptoms severity across all age groups and income groups. However, one of the limitations of this psychoeducation program is that it appears to be more advantageous for patients from low-income group as compared to middle- and high-income groups. However, this limitation can be considered a strength in a country like Pakistan where around 40% of the population lives in poverty. Usually, lower income groups tend to be worse off when it comes to treatment outcomes of any kind, but psychoeducation seems to be the avenue that appears different. Psychoeducation for schizophrenia should be explored further especially in poverty struck countries. Furthermore, the present research has opened way for an indigenous psychoeducation program for Pakistani schizophrenia patients that could potentially be used with all Urdu/Hindi speaking patients.

**Trial Registration:** **Thai Clinical Trial Registry (**TCTR20210208003).

## INTRODUCTION

Schizophrenia is a heterogeneous psychiatric syndrome which is common worldwide (Rose et al., 2010; APA, 2013). It accounts for around 14% of all the diseases contracted by the world population, and this share is increasing (Prince et al., 2007). In 2018, the World Health Organization (WHO) reported that 21 million people are suffering from schizophrenia, and these people are 2 to 3 times more likely to die earlier than the general population (WHO, 2018). In the Diagnostic and Statistical Manual of mental disorders fifth edition (DSM-5), the prevalence of schizophrenia disorder has been reported to be around 0.3% - 0.7% worldwide. Another estimate suggests that schizophrenia affects every 7 in 1000 persons aged 15-35 years (WHO, 2011). Moreover, in the United States, around 22.1% of the psychiatric population of 18 years and above has some form of schizophrenia (Sherer, 2002). Furthermore, its prevalence rate is higher in the Middle East and East-Asia when compared with that of Japan, Australia, and the United States (WHO, 2011). In Qatar, around 52.5% of the psychiatric population aged 35-49 years has schizophrenia (Ghuloum et al., 2011). In Bangladesh, the incidence is around 1.10% in adults, and around 0.10 % in children; and 2.54 of 1000 people in rural areas of the country (Rabbani et al., 2009). In India, 3 individuals out of 1000 are being affected by schizophrenia (Rabbani et al., 2009). In Pakistan, the prevalence of schizophrenia disorder is estimated to be around 1.5% in adults (Gadit et al., 2002); however, recent data regarding the prevalence of schizophrenia in Pakistan is not available.

Schizophrenia is a multifaceted disorder composed of several symptoms i.e. delusion, hallucination, disorganized speech (Huxley et al., 2014; Malaspina et al., 2014) having severe deficits (i.e. cognitive, behavioural & social) (Tandom et al., 2013), and impairments (attention, memory, executive functioning etc) (Wykes et al., 2011; Rushworth et al., 2013). Negative symptoms and comorbid disorders also appear with time when positive symptoms remain untreated (Foussias et al., 2014). Without treatment patients usually become very aggressive, raise conflicts with their family and become more illogical (Fusar-Poli et al., 2014). Long duration of illness negatively affects patients’ attitude and motivation toward treatment (Wittchen et al., 2011). Severity also affects working alliance, treatment process and treatment adherence (Wiesjahn et al., 2014). Patients with positive attitude toward treatment, respond better to medication and psychotherapy as compared to those who have negative attitude toward treatment (Sood et al., 2018). Moreover, patients with positive attitude toward treatment also start to experience positive emotions as their treatment progresses and they become more able to condone the medication side-effects (Chandra et al., 2014). Positive attitude toward treatment also makes the patients more optimistic and it enhances efficacy rate (Mohamed et al., 2009). According to an estimate, 41% - 50% patients develop negative attitude toward medication and around 70% individuals do not take medication exactly as directed in their prescription because they have a negative attitude toward treatment (Caqueo-U et al., 2015).

Impairment in motivation is one of the core deficits of schizophrenia (Nakagami, 2010). Poor motivation appears as an interior negative symptom in schizophrenia that relates with poor psychosocial and functional outcomes (Fervaha et al., 2015; Foussias et al., 2015) along with lesser treatment benefits and lesser adherence (Choi et al., 2009; Nakagami, 2008). Motivational deficits are extended in the form of negative symptoms which decrease with related interior treatment procedures (i.e. interest, drive, curiosity), initiatives (i.e. plan, pursue, engage and follow ups) and outcomes (i.e. working relation, adherence & recovery process) (Messinger et al., 2011). It is well established that motivational deficits create barriers and distort therapeutic procedures which affect decision-making and value-based rewards (Juck et al., 2006; Strauss et al., 2014) and also decreases patients’ expectations and performance regarding treatment (Bentall et al., 2010). Moreover, studies have indicated that the motivational deficits in schizophrenia affect various domains of psychological functioning (Nakagami 2010), therapeutic outcomes (Reddy 2016), cognitive functioning (Campellone 2016), adherence (Fiszdon 2016) and quality of life (Buck & Lysaker 2013). On the other hand, high motivation in patients increases the chance of resumption of normal activities (Cardeness 2013), effective response to interventions (Calton 2009), reduces apathy (Bortolon 2017) and increases the ability to develop insight about the problem (Campellone et al., 2016).

Low socioeconomic status is strongly linked with psychotic disorders (Lee at al., 2018). Multiple socioeconomic risk factors correlate with schizophrenia as they produce inequality and deprivation for the patient (Burns et al., 2014; Chan et al., 2015). Lower-income increases the chances of a person to develop schizophrenia and makes prognosis even worse (Burns et al., 2014; Saraceno et al., 1997). Positive symptoms, negative symptoms, and prevalence, all go up due to low socioeconomic level (Mazumder et al., 2015). Various studies have shown that schizophrenia disorder is more common in individuals having low-income resources (Mazumder et al., 2015), having low parental assets (Byrne, 2004), living in group homes (Wheeler 2007), and are subjected to cultural deprivation (Burns et al., 2014) and social inequality (Burns et al., 2014). People with low income not only not have enough for treatment, they also don’t have enough to fulfill their basic needs, hence they suffer more (Theodoridou et al., 2010). Pakistan is a developing country with 39.3% of its population living in some form of poverty (Haider, 2021). Hence, in Pakistan it becomes more important to seek ways that promise better outcomes for the underprivileged schizophrenia patients.

In the US, the government spent around $62.7 billion to empower health care facilities and $22.7 billion specifically for mental health in 2008, however, the situation in the low-income countries like Pakistan is quite different where only 0.4% of the total health budget was earmarked for mental health in the same year (WHO, 2009). In such a scenario, patients could easily become chronic, experience multiple episodes or may not even get any treatment (Abbas et al., 2019; Palmer et al., 2005).

The aim of the present study was to investigate the impact of a psychoeducation program, when administered on patients with schizophrenia being treated with psychotropic medications, to enhance their adherence to treatment, motivation, and lessen their symptom severity. As unmanaged/untreated symptoms of schizophrenia increase the level of severity over time (Palmer et al., 2005), which could cause severe functional impairment; patients’ motivations and attitudes toward help-seeking can play a significant role in treatment process by reducing the time between the first diagnosis and the subsequent treatment thereby improving treatment outcomes. Therefore, we made sure that our pilot study only had patients we had just started their medications (for the first time) as their first line of treatment; we took such patients because we tried to enhance their adherence to mediation as one of the goals of the psychoeducation program. To augment the treatment outcomes, we devised a psychoeducation program that was provided to all the participants enrolled in this study. In this manner, we tried to find an indigenous helpful strategy for Pakistani schizophrenia patients especially the underprivileged.

## MATERIALS AND METHODS

### Design

In this single-arm pilot clinical trial, the pre and post-assessment research design was used. Patients were recruited from hospitals and primary care clinics of Lahore, Faisalabad and Karachi(3 biggest cities of Pakistan) from June 2017 to May 2019. This information was shared with the mental health professional in order to get their referrals. During the initial screening the potential patients were instructed and guided about that how they may take part in the study. Most of the participants showed interest when the therapist gave an overview about the treatment and discussed program’s significance with the patients, nonetheless, some participants showed a lack of interest probably due to lack of awareness about the value of psychological interventions, long term engagement and/or the need for coming for frequent sessions.

### Participants

A total of 400 participants were assessed for eligibility, these 400 were those availing out-patient departments (OPDs) facilities in different psychiatric hospitals, clinics, and primary caresettings in Lahore, Faisalabad, and Karachi.. 255 participants met the study inclusion criteria and they were allocated to the interventions. Furthermore, 20 participants got excluded afterwards as they did not receive the complete allocated interventions. After follow-up, 15 participants were once again excluded due to a variety of reasons i.e., 4 participants left the city, 3 participants changed the hospital, 7 did not want to participate further in the post-assessment, and 1 respondent was referred for hospitalization due to an increase in severity of symptoms. Participants’ age range was 18-50 years (M±SD=35.45±0.27). One novel aspect of this study was that two family members of each of the participant were called in to provide guidance regarding how they can help the patients emotionally, behaviorally and socially. There were no fixed criteria for family members to be include in the psychoeducation program; any of the close family member (as per choice of the client) were contacted and instructed to take part in this program for the betterment of the patients.

Inclusion and exclusion criteria: The participants were those who were diagnosed with schizophrenia disorder according to DSM-V. Participants with duration of illness of less than 12 months and mild severity were included in the study. Participants were taken from all socioeconomic statuses with no fixed educational criteria. The exclusion criteria were as follows: 1) Patients with moderate-severe symptoms severity, 2) patients who were receiving in-patient care and/or had availed the in-patient care in the preceding year(s), 3) patients with a history of two or more previous episodes 4) patients having the duration illness of greater than 12 months, 5) patients with medical and psychiatric comorbidity, 6) patients having any kind of physical and/or intellectual disability.

### Measures

Mental Help-Seeking Attitude Scale (MHSAS; Hammer et al., 2018). The MHSAS is a self-report measure with 9-items designed to measure patients’ attitude toward treatment in terms of favorable and unfavorable outcomes. The MHSAS is scored on seven points semantic differential scale. For example, each item ranges from “useful” to “useless”. Item 2, 5, 6, 8 and 9 are negatively worded items and are reverse scored. Higher scores on MHSAS indicates that patient has positive attitude toward treatment, and s/he wants to seek help from mental health professionals while lower scores indicate that the patient has negative attitude toward treatment and is reluctant to seek help.

Patients’ Motivation for Treatment Checklist (PMTC): PMTC was prepared to assess patients’ level of motivation for treatment. The checklist was comprised of 11 items. It was completed through a detailed clinical interview. Each item was rated between “0” to “10” on a linear rating scale. A score close to 0 indicates a patient’s reluctance toward treatment (i.e., is reluctant toward getting treatment), and the ratings of 4 to 6 indicate that although the patient is reluctant toward the treatment, s/he is still willing to get the treatment anyway (i.e. delays treatment), whereas, scores close to 10 indicate that the patient is highly motivated to get the treatment (i.e., gets immediate treatment).

Positive and Negative Syndrome Scale (PANSS) by Kay et al. (1978) was administered to measure positive, negative, and general psychopathological symptoms in the participants. PANSS is comprised of 30 items and it is designed to assess symptom-severity in schizophrenia patients. PANSS has three domains: i.e., positive symptoms, negative symptoms, and general psychopathology of people suffering from schizophrenia disorder. The First 7 (i.e., P1-P7) items of PANSS have been designed to measure positive symptoms, the next 7 (i.e. N1-N7) items assess negative symptoms and the last 16 (i.e. G1-G16) items measure a patients’ general psychopathological symptoms. Each item is scored on a 7-point Likert scale (i.e. 1=abscent-7=extreme). This scale also provides symptom severity i.e., minimal, mild, and moderate-severe. This scale takes approximately 40-50 minutes to administer. In present study, PANSS was administered by a trained clinical psychologist through clinical interview.

Socio-Economic Status (SES) was assessed through patients’ family income. Information regarding patients’ financial resources was probed in a non-judgmental way during interview. On the base of given information, patients were classified into three distinct groups i.e. patients with family income of less than <50000 PKR were categorized into low-income category, patients with monthly family income between 50000-90000 PKR were considered middle-income category, and patients with monthly family income greater than 90000 PKR were considered in high-income category.

### Interventions

#### Pharmacotherapy

Antipsychotic medications; such as, “first-generation” and “second-generation” psychoanalytic has been considered an effective treatment method to deal with positive and negative symptoms of schizophrenia worldwide (Asenjo-Lobosetal., 2010; Citrome, 2013; Hartling et al., 2012). Similarly, in Pakistan, antipsychotic drugs are the first choice of treatment (Nawaz et al., 2020). Antipsychotic drugs are primarily used to treat schizophrenia disorder with the focus on managing patients’ symptoms and enabling the patients to perform their life functions (Haddad et al., 2016). After assessment and diagnosis, patients were referred to psychiatrists for medication and the psychiatrist then prescribed the medication considering patients’ level of severity and functionality. First follow-up was after one week and the next follow-ups were scheduled after every 15 days and this continued for 6 months. As it is mentioned above, antipsychotic drugs are the first choice of treatment but medication compliance is a big issue as it has been observed on other studies that around 30-40% patients respond poorly to medication because of a lack of adherence to the prescribed drug therapy (Nawaz et al., 2020). The medication non-compliance could be due to a lack of combined approach to treatment i.e., combining pharmacotherapy with psychosocial intervention, supportive therapies, rehabilitation, and awareness programs (Valencia et al., 2012). Therefore, psychoeducation program was added as a supportive intervention to enhance the pharmacotherapy treatment adherence in this study.

#### Psychoeducation program

This program was designed as an intervention that could be provided alongside the first line of treatment i.e., psychotropic medication. This psychoeducation program was based on cognitive behavior therapy to educate, guide, motivate and developing insight in patients about the problem (Valencia et al., 2012). Another focus of this program was to treat patients’ dysfunctional beliefs, cognitive distortions, and impaired motivation (Yesilyaprak et al., 2019). An overview of the psychoeducation program goals can be seen in Table 1. The psychoeducation program was designed to help the patients control their anger, distress, and elevating mood as per recommendation of Xia et al. (2011). Psychoeducation program comprised of 10 sessions with specific agenda (i.e., psychoeducation, adherence training, motivation, cognitive conceptualization, developing insight, stress management, socialization, skill training, lapse, and relapse prevention). These topics were selected from two main sources i.e., 1) from a book called ‘Cognitive Behavior Therapy-Basic and Beyond’ (Beck & Beck, 1995) 2) from material freely available from Mind’s website https://www.mind.org.uk/information-support/types-of-mental-health-problems/ in which therapist can get tailored psychoeducational strategies for their patients with schizophrenia. The psychoeducation program was compiled and then translated in Urdu language by consulting three subject experts and two language experts. Each session was conducted in a one-on-one setting with each patient. Each session frequency was planned as one session per week. At the end of this program, two sessions were conducted with the family members of each patient in which guidelines for enhancing the patient’s social and emotional functioning as per recommendations of McFarlane et al. (2003). Moreover, during these two sessions, family members of the patients were made aware of the patient’s nature of illness and a few recommendations were given to them regarding how they can effectively deal with the patients at home. A few more group activities (i.e., role playing, table discussion, sharing of life stories, music and yoga activities) were designed to enhance social interaction and social networking among the patients. Another purpose of our psychoeducational program was to reduce the level of stigma and isolatedness of the patients with schizophrenia. Pre and post testing interval was almost three months.

**Table 1.** Individual’s psychoeducation program for patients with schizophrenia disorder

| Session | Agendas | Contents |
| --- | --- | --- |
| 1 | Psychoeducation | To educate the patient about the nature of the problems. To discuss the underlying and hidden mechanisms working behind the problems. Discuss |
|  |  | the stage of illness, severity and expected future outcomes |
| 2 | Adherence training | Educate the patients about the significance of the medication. Guide and educate interactively about adherence. Plan for coping with side effects of medications and medical regimen. Formulate a daily schedule for medication and other self-care behaviors |
| 3 | Motivation and attitudes | Work with impaired motivations and attitudes. Boost motivations and positive attitude about treatment. Develop a positive attitude for treatment. |
| 3 | Cognitive conceptualization | Identification of distortions. Identification of negative schemas, thoughts, feeling and emotions, and irrational and inflexible beliefs |
| 5 |  | Evaluate and respond the automatic thoughts, and negative core beliefs. Implementation of ABC model. Cognitive restructuring. Use of alternative thoughts and beliefs. To challenge the thoughts and beliefs |
| 6 | Insight | Work with existing insight. Mechanism for developing the insight. Establish connections to develop insight through treatment and alliance |
| 7 | Stress Management | Managing stress. Managing worries. Self-help training to reduce stress. To differentiate healthy and unhealthy stress. |
| 8 | Socialization | Help and guide the client to engage in social activities and establish healthy connections with external world |
| 9 | Skill training | To provide the skill training to the client where he/she feels discomfort to perform in different areas of life |
| 10 | Relapse prevention | Educate the client, how he/she can control, manage and regulate the triggers, situations, and incidences if happens in future |
| Psychoeducation program with the family members of patients with schizophrenia disorder |  |  |
| 1 | Psychoeducation to family | To provide the understanding to family members about patient's illness and guide them, how they help the patient in difficult situation through healthy discussion, healthy dealing and avoiding stigma |
| 2 | Social and emotional support | To educate the family that how they provide social and emotional support to the patients and its significance in term of treatment |

### Procedure

All the procedures were vetted by the Institutional Review Board (IRB) of Government College University, Faisalabad. Then a researcher contacted each of the hospital authorities regarding participants’ availability and the setting. Initially, a researcher (placed in his/her respective participating hospital’s Out-Patient Department (OPD)) started with the enrollment of patients for assessment and screening. All the participants were recruited through clinical interview and standardized tests (mentioned before) in a one-on-one setting. In the first session, researcher assured the patients regarding confidentiality and that their identity will never be disclosed and they have the right to withdraw from the study at any point time if they feel like. This sample included patients who demonstrated the ability to read, write, understand and sign the informed consent form. The initial screening comprised of a diagnostic interview as well as a battery of self-report measures (discussed earlier in measures section). Participants were diagnosed according to the DSM-V criteria and then patients were assigned for the interventions. Participants were treated as per their presenting problems. Both medication and psychoeducation program were provided to them over their 10 therapy sessions and two psychoeducational sessions were provided to their family members. After 16 weeks, same battery of self-report measures was administered.

### Ethical consideration

First of all, therapist ensured the privacy and confidentiality to the participants that the received information will not be disclosed to any one that would be used only for the purpose of the research. Written informed consent was obtained from the all participants. The study protocol was approved by the Institutional Review Board (IRB) of the institution where the study was designed i.e., Government College University, Faisalabad after an in-depth scientific and technical review. The university’s Institutional Review Board makes sure that each research that is carried out must follows all the pertinent ethical guidelines and then approves a study protocol. However, as of now, the IRB of Government College University, Faisalabad does not put a compulsion of registering each clinical trial in a trial registry, the study trial was not registered at the begging stage but later on, trial was approved by the Thai Clinical Trial Registry (TCTR20210208003 with https://www.thaiclinicaltrials.org/show/TCTR20210208003)

### Evaluation of treatment acceptability

At the completion of 16-weeks psychoeducation program successfully, participants were asked to share their experience and completed the evaluation checklist that was structure to assess the quality and significance of the psychoeducation program as well as family member experience during the treatment period. Participants were asked to rate each statement on five-point rating scale from one (very poor) to five (very good). Participants shared their experience as, significance of program (very good=71%), quality of sessions (very good=65%), session timing and frequency (very good=84%), session content and understanding (very good=59%), therapist’s behavior and dealing (very good=73%), therapist maintained the privacy and confidentiality (very good=94%), involvement of family members in the treatment (very good=73%) and overall program layout (very good=83%).

### Statistical analysis

Sample size of this study was calculated through the G-Power software (version 3.1.9.7) Using a priori effect size of 0.50, α error probability of 0.001 and power of 0.99 (Faul et al., 2009). G-Power provided us with a target sample size of 138 participants. Our study has ultimately been performed with 220 participants. Descriptive statics was used to calculate sample demographic characteristics. Frequency distribution statistics was applied to check out the symptom severity on PANSS. Further, Wilcoxon’s Signed-Ranked test was used to find the difference in participants’ pre- and post-test scores on PANNSS, MHSAS, and PMTC. An alpha level of .05 was used to perform all analyses with *p-*value <.05 using IBM SPSS Statistics (Version 24).

## RESULTS

### Sample characteristics at baseline

A total of 255 participants were allocated to intervention and 235 participants received the interventions. 20 out of 235 did not completed the interventions due to variety of reasons; such as, 8 participants did not complete the therapeutic sessions, 5 participants leave during the treatment, 4 participants did not complete the post-assessment and 3 participants moved to in-patients setting. 10 therapeutic sessions were given to each patient. In sample, there were 56.08% men and 43.82% women with single 48.24%, married 38.43% and divorced/widow 13.33%. Patients’ age range was 18-52 years (M= 35.45 & SD= 10.27). All the information is given in the CONSORT flowchart (Figure. 1). Sample demographic characteristics are mentioned in the Table 2.

**Figure 1.**
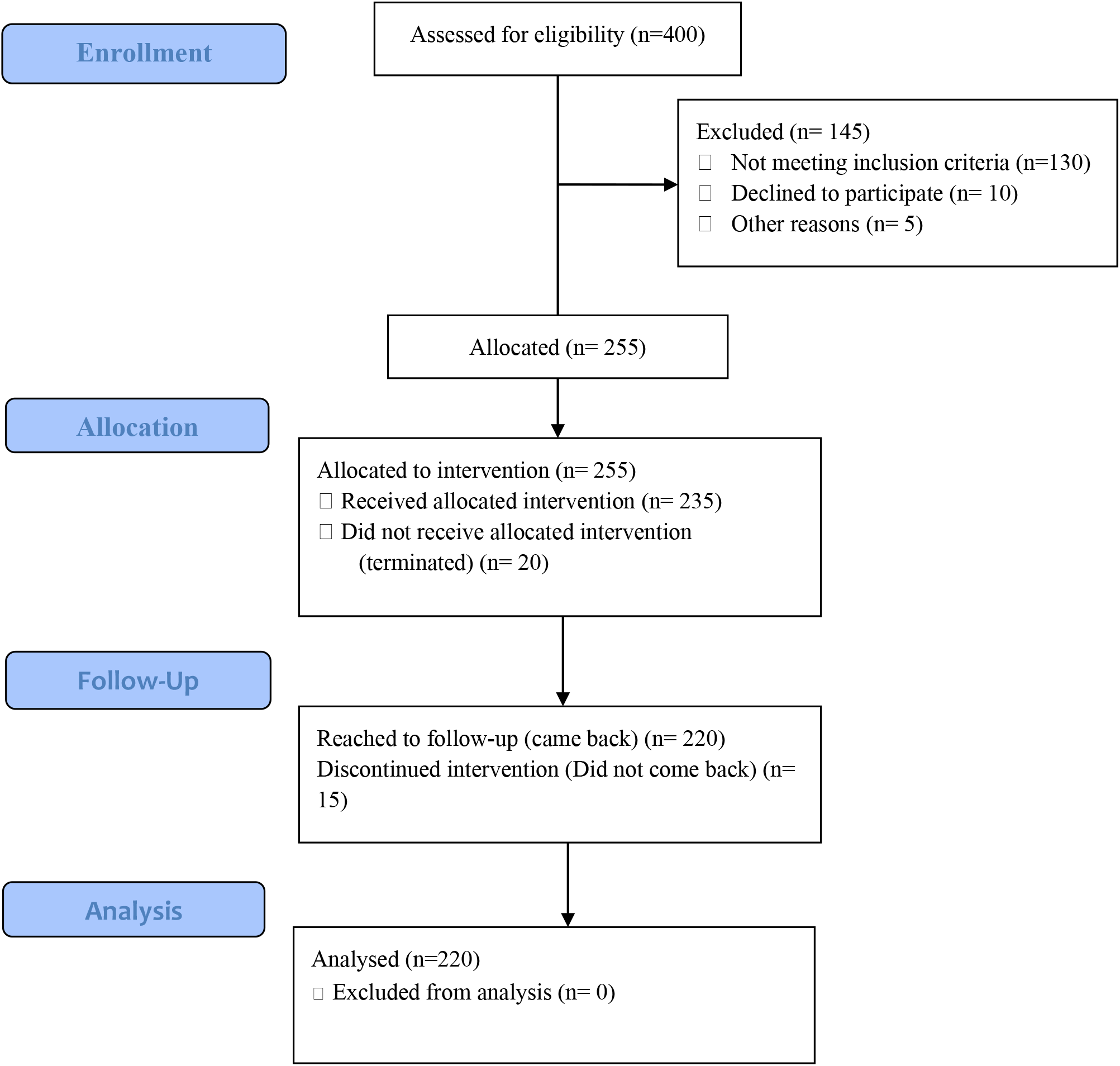
CONSORT flowchart for the (single-arm, open-label) clinical trial in patients with schizophrenia disorder.

**Table 2:** Sample demographics and baselines characteristics (n=255)

| Variables | Count (%) |
| --- | --- |
| Gender n(%) |  |
| Males | 143(56.08%) |
| Females | 112(43.82%) |
| Marital status n(%) |  |
| Single | 123(48.24%) |
| Married | 98(38.43%) |
| Divorced/widow | 34(13.33%) |
| Education n(%) |  |
| < metric | 51(20.00%) |
| Metric | 85(33.34%) |
| Intermediate | 65(25.49%) |
| Bachelor | 35(13.72%) |
| Master | 19(07.45%) |
| Family System n(%) |  |
| Nuclear | 134(52.55%) |
| Joint | 121(47.45%) |
| Monthly Income n(%) |  |
| < 50K | 81(31.77%) |
| 50K-90K | 89(34.90%) |
| 90K > | 85(33.33%) |
| Source of income n(%) |  |
| Employees | 76(29.81%) |
| Businessmen | 98(38.43%) |
| Labore | 70(27.45%) |
| Sibling support | 11(04.31%) |
| Past treatment |  |
| Medication (Only) | 196(76.86%) |
| Psychological (Only) | 21(08.23%) |
| Both treatment | 16(06.28%) |
| No treatment | 22(08.63%) |
| Duration of illness n(%) |  |
| < 6 months | 60(23.53%) |
| 6-12 months | 83(32.55%) |
| 1year | 68(26.67%) |
| 2 years | 25(09.80%) |
| 3 years & above | 19(07.45%) |
| Age M(sd) | 35.45(10.27) |

### Baseline pre-interventions assessment

After assessment and screening 235 participants suffering schizophrenia disorder were included. On PANSS 235 participants severity was determined and participants with mild severity were engaged in the psychoeducation program. Moreover, we target the participants’ motivations and attitudes toward the treatment. Participants showed scores <18 on attitude toward help-seeking were considered low level of attitude toward help-seeking attitude and participants < 5 rating on 0-10 rating scale showed low motivation toward treatment.

A Wilcoxon signed rank test revealed that significant decreased in positive symptoms severity was observed after the intervention; such as, P1 (Md = 3.34, n=220) compared to before (Md = 3.93, n=220), z = -10.80, p = .000, with a large effect size, r = .51, P2 (Md=2.49, n=220) compared to before (Md=3.14, n=220, z=-8.34, p=.000, with large effect size, r= .40, P3 (Md= 2.95, n=220) compared to before (Md= 3.75, n=220, z= -10.10, p=.000, with large effect size, r= .48, P4 (Md= 2.18,n=220) compared to before (Md= 2.60, n=220, z= -6.97, p=.000, with large effect size, r= .33, P5 (Md= 3.28,n=220) compared to before (Md= 3.66, n=220, z= -6.25, p=.000, with large effect size, r=29, P6 (Md= 3.38, n=220) compared to before (Md=3.96, n=220, z= -8.67, p=.000, with large effect size, r= .41 and P7 (Md= 3.43, n=220) compared to before (Md= 3.95, n=220, z= -7.04, p=.000, with large effect size, r= .34. Similarly, significant symptoms severity decreased after interventions on negative symptoms; such as, N1 (Md= 1.79, n=220) compared to before (Md= 2.43, n=220, z= -8.91, p=.000, with large effect size, r=42, N2 (Md= 2.45, n=220) compared to before (Md= 2.74, n=220, z= -6.31, p=.000, with large effect size, r= .30, N3 (Md= 2.50, n=220) compared to before (Md= 2.70, n=220, z= -6.63, p=.000, with large effect size, r= .32, N4 (Md= 2.21, n=220) compared to before (Md= 2.27, n=220, z= -2.08, p=.038, with large effect size, r= .10, N5 (Md= 2.15, n=220) compared to before (Md= 2.40, n=220, z= -6.52, p=.000, with large effect size, r= .31, N6 (Md= 2.51, n=220) compared to before (Md= 2.59, n=220, z= -2.09, p=.036, with large effect size, r= .10 and insignificant difference was found on N7 (Md= 2.12, n=220) compared to before (Md= 2.31, n=220, z= -1.80, p=.072, with lower effect size, r= .09. Moreover, significant change was observed on general psychopathological symptoms severity after interventions on G1 (Md= 2.30, n=220) compared to before (Md= 2.71, n=220, z= -8.88, p=.000, with large effect size, r= .42, G2 (Md= 2.69, n=220) compared to before (Md= 3.10, n=220, z= -6.94, p=.000, with large effect size, r= .33, G3 (Md= 2.65, n=220) compared to before (Md= 2.90, n=220, z= -7.48, p=.000, with large effect size, r= .36, G4 (Md= 2.37,n=220) compared to before (Md= 2.82, n=220, z= -7.42, p=.000, with large effect size, r= .35, G6 (Md= 2.54, n=220) compared to before (Md= 3.06, n=220, z= -10.24, p=.000, with large effect size, r= .49, G7 (Md= 1.68, n=220) compared to before (Md= 1.80, n=220, z= -5.00, p=.000, with large effect size, r= .24, G8 (Md= 1.56, n=220) compared to before (Md= 1.71, n=220, z= -5.83, p=.000, with large effect size, r= .28, G11 (Md= 1.66, n=220) compared to before (Md=2.04, n=220, z= -7.03, p=.000, with large effect size, r= .34, G12 (Md= 2.41, n=220) compared to before (Md= 3.03, n=220, z= -11.08, p=.000, with large effect size, r= .53, G13 (Md= 3.02, n=220) compared to before (Md= 3.85, n=220, z= -9.68, p=.000, with large effect size, r= .46, G14 (Md= 2.72, n=220) compared to before (Md= 3.08, n=220, z= -8.84, p=.000, with large effect size, r= .42, G15 (Md= 2.55, n=220) compared to before (Md= 2.94, n=220, z= -7.70, p=.000, with large effect size, r= .37 and insignificant difference was found on G5 (Md= 1.88, n=220) compared to before (Md= 1.96, n=220, z= -0.75, p=.452, with lower effect size, r= .04, G9 (Md= 2.44, n=220) compared to before (Md= 2.46, n=220, z= -1.42, p=.157, with lower effect size, r= .07, G10 (Md= 2.55, n=220) compared to before (Md= 2.58, n=220, z= -1.61, p=.109, with lower effect size, r= .08, and G16 (Md= 3.31, n=220) compared to before (Md= 3.35, n=220, z= -1.43, p=.154, with l0wer effect size, r= .07 respectively (Table 3).

**Table 3.**
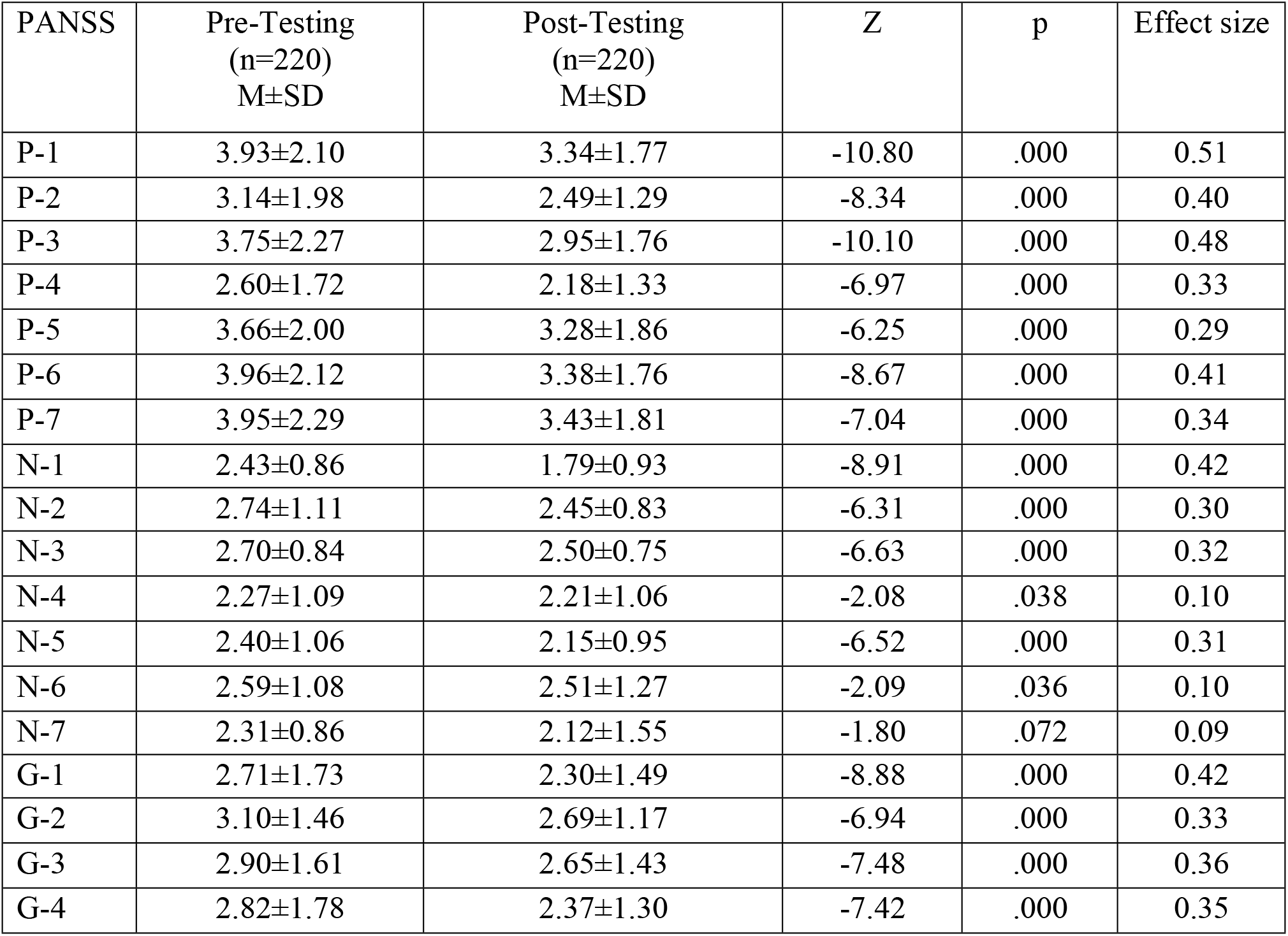
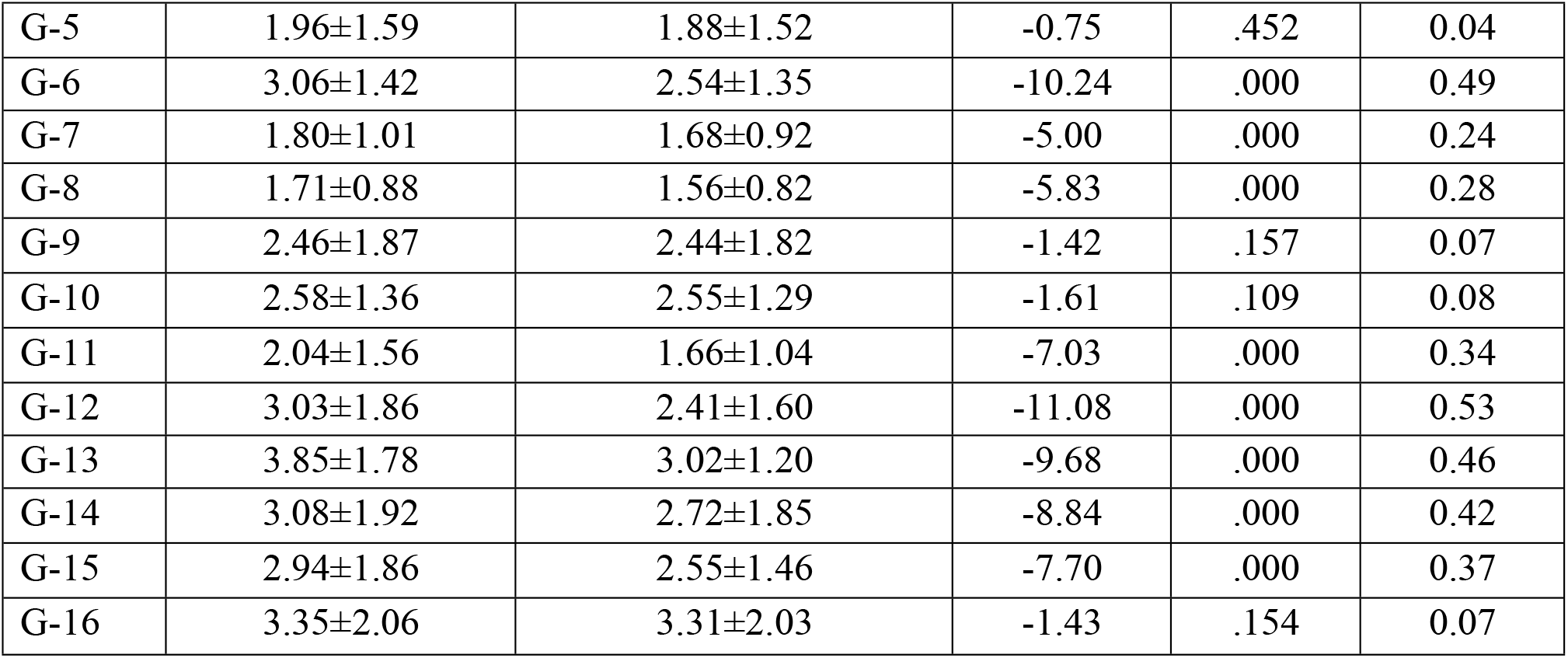
Wilcoxon’s Signed-Ranked test between pre and post-testing on PANSS.

A Wilcoxon-signed rank test revealed that significant change in symptoms severity was observed after 3 month psycho-education program among patients with schizophrenia disorder; such as, positive symptoms (Md=21.05, n=220) compared to before (Md=25.00, n=220, z=-12.47, p=.000, with large effect size, r= .59, negative symptoms (Md=15.74, n=220) compared to before (Md=17.44, n=220, z=-9.52, p=.000, with large effect size, r= .45, and general psychopathological symptoms (Md=38.32, n=220) compared to before (Md=43.40, n=220, z=-12.72, p=.000, with large effect size, r= .61. Moreover, findings indicate that 3 months psychoeducation program significantly improved patients’ motivation and attitude toward treatment; such as, HSAT (Md=39.03, n=220) compared to before (Md=28.27, n=220, z=-10.43, p=.000, with large effect size, r= .50, and PMFT (Md=5.69, n=220) compared to before (Md=4.85, n=220, z=-12.43, p=.000, with large effect size, r= .59 respectively (Table 4).

**Table 4.** Wilcoxon’s Signed-Ranked test between pre and post-testing on PANSS domains, help-seeking attitude, and motivation toward treatment.

| PANSS | Pre-Testing<br>(n=220)<br>M±SD | Post-Testing<br>(n=220)<br>M±SD | Z | p | $\eta_p^2$ |
| --- | --- | --- | --- | --- | --- |
| PANSS |  |  |  |  |  |
| Positive symptoms | 25.00±8.89 | 21.05±6.91 | -12.47 | .000 | .59 |
| Negative symptoms | 17.44±5.19 | 15.74±5.33 | -9.52 | .000 | .45 |
| General symptoms | 43.40±10.73 | 38.32±9.25 | -12.72 | .000 | .61 |
| HSAT | 28.27±12.89 | 39.03±13.27 | -10.43 | .000 | .50 |
| PMFT | 4.85±1.98 | 5.69±2.05 | -12.43 | .000 | .59 |

*Note: $\eta_p^2$ = Partial Eta Squared, PANSS= Positive and Negative Symptoms Severity Scale, HSAT= Help-Seeking Attitude Toward Treatment, PMFT= Patient's Motivation for Treatment*

Findings show (Table 5) that psychoeducation programs substantially improve patients’ motivation after 3-months. For example, in pre-treatment, patients with low motivation were 103(46.82%), and in post-treatment, there were 48(21.81%), which indicates 55(25.00%) participants improved their motivational levels. During pretreatment, patients with moderate motivation were 64(29.10%), and post-treatment, there 74(33.64%), which reflects that 10(4.55%) participants improved their level of motivation. Similarly, at the pretreatment level, patients with high motivational levels were 53(24.08%), and at post-treatment, there were 98(44.55%), which shows 45(20.45%) patients improved their high motivational levels.

**Table 5.** Motivational level on rating scale (0-10) from pretreatment to posttreatment (N=220)

| Motivational level | Pretreatment<br>n(%) | Post treatment<br>n(%) | Change %<br>(Pre – Post)<br>n(%) |
| --- | --- | --- | --- |
| Low motivation (<4) | 103(46.82%) | 48(21.81%) | -55(25.00%) |
| Moderate motivation (4-6) | 64(29.10%) | 74(33.64%) | 10(4.55%) |
| High motivation (7-10) | 53(24.08%) | 98(44.55%) | 45(20.45%) |

Results (Figure 2) indicate that the psychoeducation program substantially decreased their positive, negative, and general symptoms after 16 weeks of sessions. A significant decrease in positive symptoms was observed; similarly, change was noticed in negative and general psychopathological symptoms. This reflects that the psychoeducation program effectively developed insight and awareness about the symptoms among patients.

**Figure 2:**
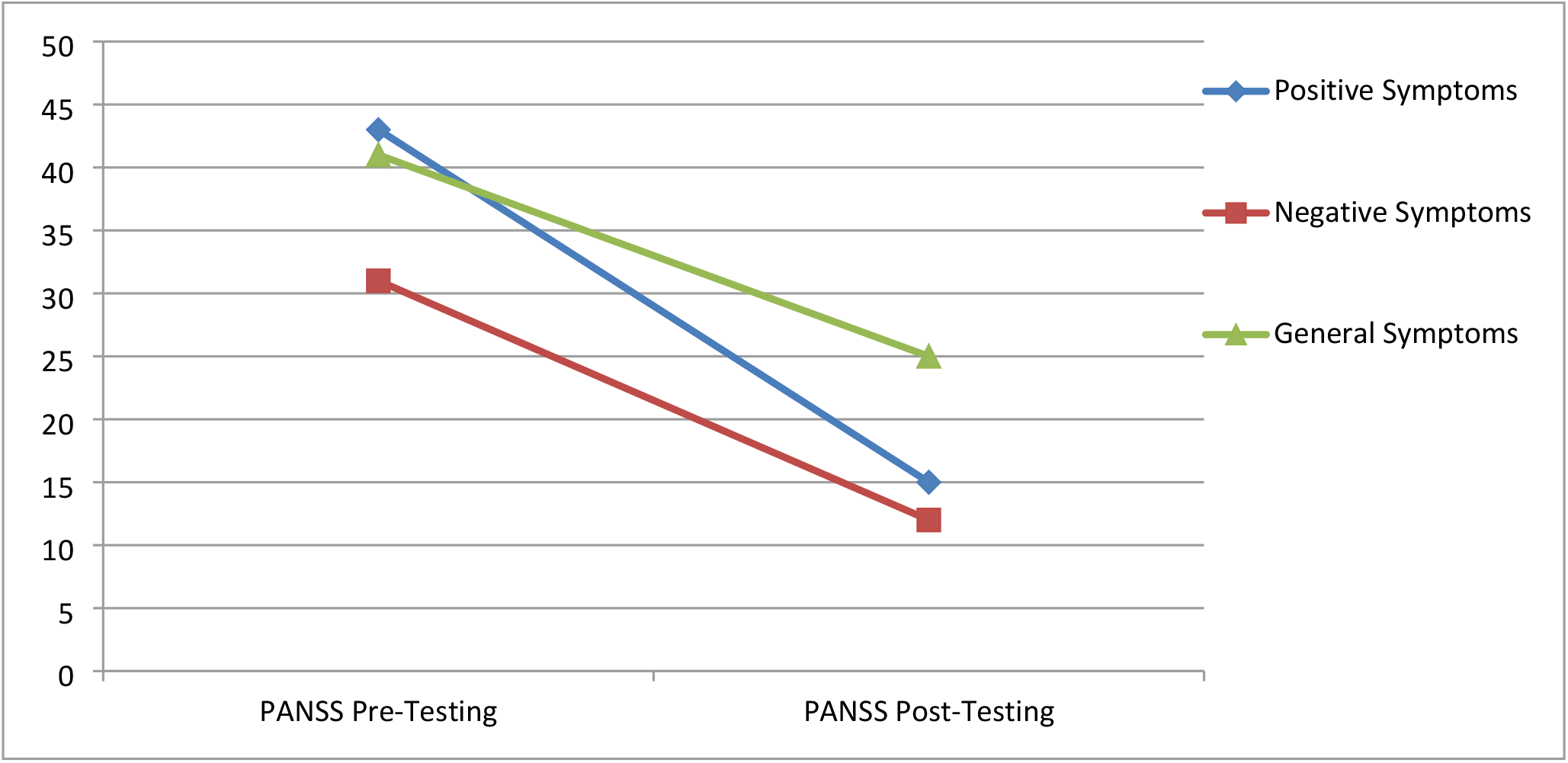
The pre- and post- assessment Results of PANSS after 16 weeks of sessions of patients with Schizophrenia Disorder.

Findings (Figure 3) shows that the psychoeducation program was an effective intervention to address positive, negative, and general symptoms severity among patients of all socioeconomic statuses. Furthermore, it was observed that patients in the low-income group had more symptoms severity compared to middle- and high-income groups. However, the symptom severity of low-income group also got reduced the most as a possible effect of the psychoeducation program.

**Figure 3:**
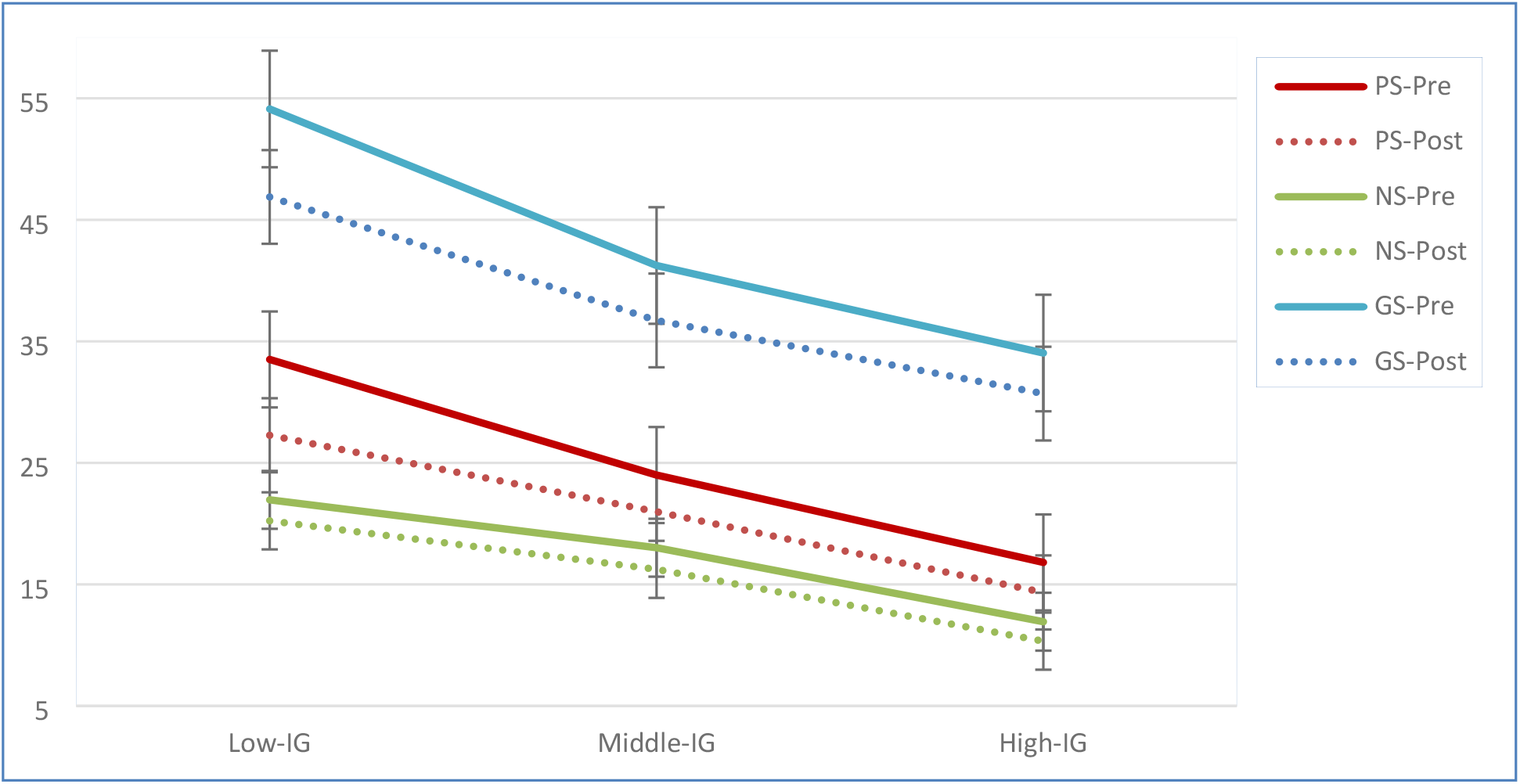
Symptoms change between pre- and post-assessment among patients with low, middle and high socioeconomic status.

Furthermore, findings reveal that there was no big difference between age groups on symptoms severity. Furthermore, after the psychoeducation program, a significant reduction can be seen in positive, negative and general symptoms severity among patients of all age groups (Figure 4).

**Figure 4:**
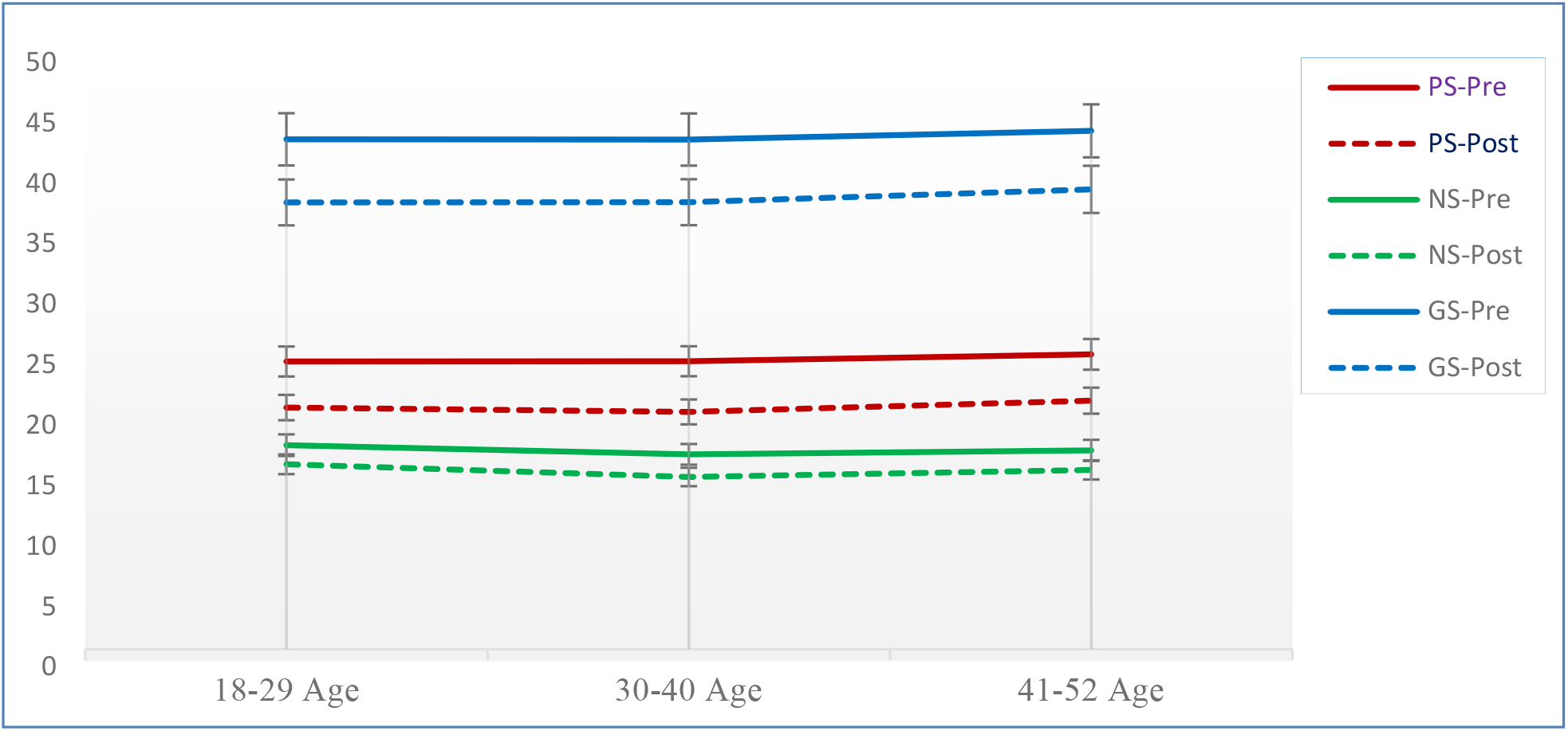
Impact of psychoeducation program on age and symptoms severity using pre and post- assessment outcomes

## DISCUSSION

Our study findings reported psychoeducation program produced substantial improvement. This 16-week psychoeducation program was planned to address patients’ positive, negative, and general symptoms and improve patients’ motivation and help-seeking attitude toward treatment. This piloting clinical trial was the first time we planned in Pakistan after reviewing patients’ overall low progress on treatment with no significant change in symptoms severity. It was observed that patients’ help-seeking attitude toward treatment, motivations and income groups are the major hindrance to treatment efficacy. To address these factors, the psychoeducation program was structured. This psychoeducation program significantly addresses the severity of positive, negative and general psychopathological symptoms (Leucht et al., 2013). This psychoeducation program created insight and awareness among patients to manage their symptoms. Adherence training helped the patients take medication properly and cope with the stressors and emotional disturbances using skill training. The therapist also explained how emotional disturbance causes social, cognitive, and behavioral dysfunctions (Carter & Barch, 2007; Sullivan et al., 2012).

Some evidence support that a psychoeducation program is an effective approach to addressing and managing positive and negative symptoms severity (Xia, 2011; Chien et al., 2013). Another piece of evidence reported without a psychoeducation program is that the psychotic episode, prodromal period, and occasional symptoms enhance symptoms severity. Eventually, symptoms transit into negative symptoms and collectively produce severe dysfunctional outcomes (Sabbag et al., 2011). Early symptoms management empowers patients’ behavioral and cognition functioning and the chance of successful recovery (Brown et al., 2012, Hoe et al., 2012; Fitch et al., 2010, Frith & Frith, 2012). Psychotropic mediation is the better choice of treatment (Asenjolobos et al., 2010; Hartling et al., 2013), and it produces substantial improvement along with a psychoeducation program as a supportive intervention (Kalkstein et al., 2010; Naber & Lambert, 2009).

Furthermore, it can be noted from our study that our psychoeducation program benefits most of the age-groups almost equally. This finding is consistent with the findings of McFarlane, et al. (1995) who also saw similar trends in patients from 18 to 45 years of age. Another factor that may have been behind this finding could be that the urgency of treatment is more important (Malik, et al., 2010).

Our results indicate the psychoeducation program significantly changed patients’ motivation after 16-weeks sessions. The program enhanced patients’ motivation which usually helps the person engage in treatment and be concerned with treatment procedures. Patients with adequate motivation pursue treatment properly compared to those with low or inadequate motivation. In our culture, patients commonly believe in medication, having no awareness and particular knowledge about the future consequences and never considering the role of self-attitude and motivation. Nakagami et al. (2010) identified inadequate motivations as the main reasons for non-compliance and distress cycle when patients avoid treatment, use irregular medication, and have no insight into the problem. This usually leads to disturbed reality contact, attitudes toward life, and motivations (Choi et al., 2010). High motivation toward treatment plays an essential role in treatment factors and functional outcomes (Horan et al., 2006). This psychoeducation program boosted the patients’ motivation toward treatment, making them optimistic and eagerly engaged in the treatment process (Medalia & Saperstein, 2011; Sorokin et al., 2017). High motivations could urge a patient internally to move toward a better life. In contrast, less motivated patients have negative treatment beliefs, i.e., they deem it worthless and express increased cognitive distortions (Kessler et al., 2001). In addition, less motivated patients sustain their positive and negative symptoms throughout their illness (Bentall et al., 2010; Messinger et al., 2011; Reddy, 2016; Medalia et al., 2010).

Another factor that plays an important role is the patient’s help-seeking attitude toward treatment. Psychoeducation programs change patients’ attitudes by providing education, insight, stress management, skill training, and motivation (Mohamed et al., 2014). This program remained effective in promoting a positive attitude toward treatment, and it also enhanced treatment adherence (Ram et al., 2019). It is observed that when patients feel compliance and satisfaction with the treatment, they usually develop a positive help-seeking attitude. Therefore, this psychoeducation program produced effective outcomes by providing guidance, insight, and therapeutic support to reduce the symptoms’ severity. Moreover, patients’ positive help-seeking attitude may be promoted by receiving therapeutic guidance and training, which develops insight in a patient toward their problem. A negative help-seeking attitude toward treatment creates negative beliefs in patients’ minds that would cause treatment avoidance and follow-ups (Ram et al., 2019).

Besides motivational, attitude, and attributional factors, the present study found quite high differences between high-income group, middle-, and low-income groups on pretest of symptom severity, however all three groups benefitted from our psychoeducation program and their symptom severity lessened. An interesting thing to note here is that, although symptom severity was highest in the low-income group (on pretest) but this group also benefitted the most from our psychoeducation program and got highest reduction in their symptom severity (posttest). Most of the studies report that underprivileged are usually worst off if they get afflicted with schizophrenia. Nakagami et al., (2010) found financial obstacles and access to treatment problem as significant factors causing treatment non-compliance in Indian sample of patients with schizophrenia. Somewhat similar findings were identified by Burns et al., (2014), Chan et al.,(2015) and Gururaj et al., (2005), that various countries with higher income-inequality, there is comparatively higher prevalence of schizophrenia. Whereas on the other hand, people with high income resources, respond better to treatment (Citome et al., 2013). In fact, patients from low socioeconomic background already have very limited resources to manage their families’ daily needs; additionally, there is absence of facilities like medical insurance or free treatment. So having a psychiatric patient in a family adds burden on the family’s already degraded resources (Millan et al., 2014). On the other side, patients with high income resources can avail better treatment opportunities; they can afford better practitioners, bear medication expenses and may have life without extreme debt burdens (Zagozdzon et al., 2017). Moreover, patients with high income status usually approach practitioner on right time because they have no issue of affordability while patients with low-income resources prefer local cheap remedies, quack practitioners, and faith healers to manage the symptoms, thus this delay adversely effects their symptoms severity. Beside these, another reason is a lack of treatment facilities, and support system which also causes increases in the severity in patients over time (Medalia & Choi, 2009). However, one aspect that may make this pilot study worth expanding is that the low-income group responded better to our psychoeducation program than middle income and high-income groups. It could be due to the attention and focus that the underprivileged patients got from our psychoeducation program that they otherwise do not get a chance as most of their family members are busy earning for survival.

## Conclusion

It is concluded that the management of patients with schizophrenia disorder using our psychoeducation program (alongside medication) reduces the frequency of positive, negative, and general psychopathological symptoms across most age groups and economic conditions. Patients’ motivation and positive attitude toward seeking help is very important to sustain patients’ engagement in treatment so our psychoeducation program has these factors built into it. Socioeconomic status is another cause which significantly increases or decreases the process and outcomes of the treatment; our psychoeducation program benefits all income groups of patients, however, it tends to benefit the underprivileged patients more. Finally, psychoeducation program (alongside medications) increases treatment efficacy and supports the process of recovery.

## Limitations of the study

The current study has some limitations. The limitation built into this pilot study is that it is a single arm pilot study of patients with schizophrenia who were already on medication, so, our psychoeducation program cannot be disentangled from the psychoactive medications the patients were receiving as there is no control group. Furthermore, only patients who were at the initial stage of their illness and having relatively less severity, were targeted for this research for different outpatient settings. Moreover, major focus was given to patients’ level of motivation and attitude toward treatment and other factors were not targeted that might play helping role in the treatment, such as, patients’ social and emotional support system, role of cognitive functioning, emotional intelligence, well-being, and others associated factors. Another factor is that the psychoeducation program was mainly designed to develop insight, enhance adherence, increase motivation, and bring about positive attitudes in the patients toward treatment and was not a comprehensive psychotherapeutic treatment intervention in itself. Extensive psychotherapy along with medication could play an even more effective role in the recovery process.

### The implication of the study

This pilot study investigated the impact of a psychoeducation program working through motivation and attitude to address symptoms severity. This program produced significant positive advancements in addressing symptoms severity and also provided a road map to work on and explore further options for psychoeducation programs to be conducted with Pakistani patients. For this reason, this study is unique in nature and it could facilitate practitioners and clinicians working in a primary care setting of Pakistan. This study can also help mental health professional understand the importance of concurrent initiation of psychological support alongside psychotropic medications. Moreover, the current study highlights that the reluctance of patients with schizophrenia to participate in treatment and low motivation and negative help-seeking attitudes toward treatment are some of the problems amenable to psychoeducation. Considering this viewpoint, if we work on patients’ motivation and attitude, we may engage the patients in treatment and produce better treatment outcomes. Our psychoeducation program can be used with different psychiatric patients where a lack of insight and motivation for treatment is felt by the clinician. Our pilot study has provided the basis of a workable psychoeducation program in Urdu, future researchers could use our program to further conuct full-scale randomized control trials.

## Data Availability

If you required it would be upload as per your direction and need

## DISCLOSURE

None

## CONFLICT OF INTEREST

None

## FUNDING

None

## Notes

### Competing Interest Statement

The authors have declared no competing interest.

### Clinical Trial

Thai Clinical Trial Registry (TCTR20210208003 with https://www.thaiclinicaltrials.org/show/TCTR20210208003)

### Clinical Protocols

https://www.thaiclinicaltrials.org/show/TCTR20210208003

### Funding Statement

The author(s) received no specific funding for this work.

### Author Declarations

The study protocol was approved by the Institutional Review Board (IRB) of the institution where the study was designed i.e., Government College University, Faisalabad after an in-depth scientific and technical review.

